# Impact of increased funding for Stop Smoking Services in England on quit attempts: population-based study 2021-2025

**DOI:** 10.64898/2026.06.29.26356662

**Authors:** Vera Helen Buss, Lion Shahab, Linda Bauld, Susan Michie, Jamie Brown

**Affiliations:** Department of Behavioural Science and Health, University College London, London, UK; Behavioural Research UK, Edinburgh, UK; Usher Institute, University of Edinburgh, UK; Centre for Behaviour Change, University College London, UK

**Keywords:** Health Policy, Tobacco Smoking, Smoking Cessation, Interrupted Time Series Analysis, England

## Abstract

**Background:** The UK Government aims to reduce smoking rates by implementing new, and investing in existing, tobacco control strategies including increased funding for Stop Smoking Services (SSS) in England. This study examined whether the additional funding starting in April 2024 was associated with a detectable increase in quit attempts supported by SSS and whether it was cost-effective.

**Methods:** We used data from the Smoking Toolkit Study, a repeat cross-sectional survey conducted in 2021 to 2025. Adults aged ≥18 years who smoked cigarettes and had made a quit attempt in the past year were included (n_weighted_=5,076). The outcome was monthly prevalence of past-year quit attempts supported by SSS. We fitted general additive models with a step change in April 2024 to represent the start of the increased funding. We adjusted for tobacco tax increases, the Swap-to-Stop scheme, age, gender, and a measure of socioeconomic position. In an unplanned analysis, we extended the time series back to 2006. For the cost-effectiveness, we estimated incremental cost-effectiveness ratios for the total population and age groups, accounting for future lifetime cessation.

**Results:** In the primary model, the April 2024 step change was not statistically significant (adjusted odds ratio: 1.13; 95% CI: 0.52, 2.49). The cost-effectiveness analysis ranged from cost-effective to extremely ineffective (incremental cost-effectiveness ratio (ICER): £104,126, 95% CI: £939,398 to £8,293). When using the extended time series, the adjusted odds ratio for the step change was 2.70 (95% CI: 2.03, 3.60) and the intervention was cost-effective (ICER: £13,857; £21,393 to £9,620).

**Conclusions:** Compared with the long-term trend, increased funding to SSS in England in 2024 appeared to lead to an increase in quit attempts supported by SSS at the population level. This result is somewhat uncertain because our primary pre-planned analyses assessing the impact relative to a more recent trend were insensitive.

**KEY MESSAGES:**

- **What is already known on this topic** – SSS are effective in helping people quit smoking. However, until recently, substantial funding cuts in England by the UK Government had reduced service capacity and contributed to low usage during quit attempts.
- **What this study adds** – The renewed investment in SSS introduced in April 2024 may be associated with an increase in the number of quit attempts supported by these services in England, depending on the time horizon chosen.
- **How this study might affect research, practice or policy** – Together with existing evidence and reported increases in service throughput, these results strengthen the rationale for sustained funding in SSS as part of England’s comprehensive tobacco control strategy.

## INTRODUCTION

Cigarette smoking is the leading behavioural risk factor for premature death and illness as well as health inequalities (1, 2). The UK Government aims to reduce smoking rates by implementing new, and investing in existing, tobacco control strategies, including increasing the funding for Stop Smoking Services (SSS) in England (3). This study assessed its potential impact on quit attempts and its cost-effectiveness.

SSS provide support to people who want to quit smoking (4). Services are provided locally by trained staff and can include group therapy or one-to-one support. People can also get smoking cessation medication through these services, either for free or a small standard prescription charge (5). People are more likely to quit smoking successfully if they use support, including SSS (6–8). A study from 2012-2013 found that the carbon monoxide-validated quit rate among clients of SSS in England was 8% after one year, meaning that over 18,000 premature deaths were prevented by the services that year (8). The study also reported that the quit success rate was higher when support came from specialist stop smoking advisors than from other healthcare professionals such as pharmacists or general practitioners. However, uptake of these services is relatively low: Less than 5% of those who attempted to quit smoking in the past year in England reported having used such a service, and many who try to quit do not use any aids at all (6).

In England, the funding for the services is provided by the Department of Health and Social Care and allocated to local authorities (9). To increase smoking cessation efforts, the UK Government has more than doubled its spending on SSS in England by adding a further £70 million per year, bringing the total to £138 million per financial year from 2024-2025, starting in April 2024, until at least 2028-2029 (10). Funding is allocated annually to local authorities based on their local smoking rates, using the average smoking prevalence over a 3-year period. Local authorities have some flexibility to allocate small portions of the funding for wider tobacco control or measures to reduce youth vaping. However, the majority should be spent on SSS (10).

The National Health Service England publishes quarterly statistics on SSS, including the number of people setting a quit date (11). Service provider data suggests that there has been an approximately 20% increase in the number people setting a quit date between the financial years 2023-2024 and 2024-2025, with an indication of a further increase in 2025-2026 (11). However, there is a risk that these figures overestimate the number of people making SSS-supported quit attempts because service provision could also be seen as performance indicator and thus provide an incentive for service providers to misclassify clients as having set quit dates (12). Therefore, it is helpful to use an independent data source for the analysis. With the present study, we assessed the potential effect of the additional funding on SSS-supported quit attempts as well as its cost-effectiveness, using data from a representative population-based survey. The research questions (RQ) were:

1. Has there been a detectable change in quit attempts supported by SSS among adults who tried to quit smoking in the past year in England since April 2024 (i.e., start of financial year 2024/2025) when the additional funding for these services started?
2. How cost-effective was the additional funding in the financial year 2024-2025 (i.e., April 2024 to March 2025) in terms of cost per life-year gained?

## METHODS

### Study design and participants

We used data collected between October 2021 and October 2025 as part of the Smoking Toolkit Study (STS), a population-based, repeat cross-sectional survey (13). Ipsos, a market research company, conducted monthly telephone interviews. Each wave included roughly 1,700 adults aged ≥18 years in England. Households were sampled using a hybrid of random location and quota sampling, and one member per household was interviewed until quotas based on the probability of being at home (e.g., age, gender, working status) were met (13). The research team received deidentified data. The manuscript followed the Strengthening the Reporting of Observational Studies in Epidemiology (STROBE) statement (14) and the protocol was pre-registered on the Open Science Framework (https://osf.io/r59eu).

In the primary analysis, we included adults aged ≥18 years who smoked cigarettes and had attempted to quit in the past year. We also conducted a sensitivity analysis, which included all who had smoked cigarettes in the past year (i.e., people currently smoking cigarettes as well as those who had stopped within the past 12 months – not just those reporting a quit attempt). Smoking status was determined by the question “Which of the following best applies to you?”. Responses “I smoke cigarettes (including hand-rolled) every day”, “I smoke cigarettes (including hand-rolled), but not every day”, or “I have stopped smoking completely in the last year” classified participants as having smoked cigarettes in the past year. Quit attempts were identified by reporting at least one attempt. We focussed on those who smoked cigarettes because SSS are focused on cigarette smoking cessation. It is possible that some of those who quit in the past year smoked other tobacco products; we cannot differentiate this in our dataset, but less than 2% people currently smoking use non-cigarette tobacco.

### Dependent variable

Participants were asked “Which, if anything, of the following did you try to help you stop smoking during the most recent serious quit attempt?”. Those selecting “Attended a Stop Smoking group” or “Attended one or more Stop Smoking one-to-one counselling\advice\support session\s” were classified as having made past-year quit attempts supported by SSS. Monthly prevalence was calculated as the proportion of people making SSS-supported quit attempts among all who tried to quit in the past year.

### Sensitivity analyses

We expanded the outcome to include prescription medication (“Nicotine replacement product on prescription or given to you by a health professional”, “Zyban (bupropion)”, “Champix (varenicline)”, or “Cytisine (e.g. Tabex, Tactizen or Desmoxan)”) as these are often provided via SSS (alternatively, these medications can be accessed via general practitioners or community pharmacies (15, 16)).

Figure S1 shows weighted percentages of quit attempts supported by SSS or different types of prescription medication among people who smoked cigarettes in the past year. During the study period (2021-2025), nicotine replacement products on prescription were most popular. Varenicline was temporarily withdrawn in the UK from October 2021 and was reintroduced in August 2024 (17). However, use has remained low since its reintroduction (Figure S1).

In another sensitivity analysis, we changed the denominator to assess the proportion of people making SSS-supported quit attempts among all who smoked cigarettes in the past year (not only those who tried to quit).

We also intended to conduct a sensitivity analysis in which we restricted the outcome to quit attempts made in the past month; however, the absolute numbers were so low that we were unable to perform this analysis.

### Independent variables

Time was coded by survey wave (1 = October 2021 to 49 = October 2025). A step-change variable indicated the intervention period (0 = pre-intervention from October 2021 to March 2024; 1 = intervention from April 2024 to October 2025), reflecting additional SSS funding from April 2024.

### Sensitivity analyses

We also tested an early effect pre-specified as starting in February 2024, as it is common practice in the public sector to increase spending toward the end of the financial year to avoid underspending (18), and the impact of public health measures sometimes begins when they are announced rather than only upon their official commencement (19, 20). Additionally, we tested a delayed start pre-specified in July 2024 and a change in trend, as there could have been a gradual change instead of an abrupt one.

Seasonality was adjusted using a cyclic cubic spline (21) for month (ranging 1-12). Other covariates included age, gender, and occupational social grade as a measure of socioeconomic position. Age was included as a continuous variable modelled using restricted cubic splines (22) for RQ1, and as a categorical variable (18-34, 35-44, 45-54, 55-64, ≥65) for RQ2. Gender was included as binary (women/men); non-binary was included in descriptive statistics only due to small numbers. Occupational social grade was dichotomised (ABC1 = more advantaged; C2DE = less advantaged) (23).

Analyses adjusted for tobacco tax increases (dummy = 1 for November each year and April 2023; otherwise 0) (24, 25) and England’s Swap-to-Stop programme (dummy = 1 from December 2023 onwards), a smoking cessation intervention providing free vapes and behavioural support (26).

### Analysis

We conducted the analysis in RStudio (version 2022.07.2, R version 4.2.1), using a significance level of 0.05 and reporting 95% confidence intervals (CI). Missing values for variables are listed in supplementary Table S1. We performed a complete case analysis. For 1.8% (n=309) who reported age only as a range, we imputed the median for that range. Data were weighted using raking (27) to match the population of England.

### Change in quit attempts supported by SSS

We fitted a general additive model:

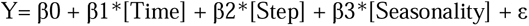

Where:

- Y = rate of quit attempts supported by SSS
- β0 = baseline average
- β1 = time trend
- β2 = intervention effect (pre/post)
- β3 = seasonal adjustment using cyclic cubic spline

We also ran adjusted models, including above-inflation tax increases, Swap-to-Stop, age, gender, and social grade.

### Sensitivity analyses

We included six sensitivity analyses testing variations in the intervention effect, outcome, and population. Analysis 7 was not specified in the protocol; it was included to increase the power with a longer pre-intervention period following insensitive results on the primary analysis.

1. Step change in February 2024 (early effect)
2. Step change in July 2024 (delayed effect)
3. Interaction between time and step change (trend change post-intervention)
4. Outcome: quit attempt supported by SSS including prescription medication
5. Denominator: all who smoked cigarettes in past-year
6. Time series starting in November 2006 (i.e., first data available in STS)

### Cost-effectiveness

We used a model by Stapleton and West (see (28) for further details), previously applied to STS data (e.g., (29)), to assess the cost-effectiveness of the additional funding based on the odds ratio for the step change (i.e., the difference in the odds of SSS-supported quit attempts with and without the additional funding in April 2024) . We estimated incremental cost-effectiveness ratios for the total population and five distinct age groups (18-35, 35-44, 45-54, 55-64, ≥65). The incremental cost-effectiveness ratios were derived as the additional funding per person smoking divided by the attributable discounted life-years (DLY) gained per person due to the additional funding. The model accounted for future lifetime cessation (life-years gained per group: 1.084, 1.588, 1.577, 1.161 and no gain for quitting ≥65 years (28)). Population estimates were based on the 2024 annual average cigarette smoking prevalence from the STS and the Census age distribution (30).

We assumed that 2.5% of the reported quit attempts result in permanent success (29, 31). We assessed the intervention effectiveness overall and for each age group using the primary model (adjusted for seasonality, above-inflation tax increases, Swap-to-Stop) and the long-term series starting in 2006 (adjusted for seasonality only). To calculate 95% CIs, we took 10,000 draws of the model coefficients and, for each draw, generated predicted probabilities for effectiveness with and without additional funding. These were transformed to probabilities and differenced to produce a simulated distribution of percentage-point effects, from which the 2.5^th^ and 97.5^th^ percentiles were taken as the 95% CIs. For health-gain outcomes, we assumed that additional funding would not plausibly result in negative attributable DLY gained per person who smokes; therefore, simulated DLY values below zero were truncated at zero, and non-finite incremental cost-effectiveness ratio (ICER) values arising from near-zero DLY gains were removed prior to calculating ICER confidence intervals. Details on the calculations and the underlying assumptions are provided in supplementary Tables S9-10.

## RESULTS

We included 12,314 participants (unweighted) with complete data who smoked cigarettes in the past year. In the financial year 2021/22 (i.e., October 2021 to March 2022), 0.5% of them made a past-year quit attempt supported by SSS and 1.2% of those who tried to quit in the past year. These figures increased to 1.8% and 4.8%, respectively, in the financial year 2025/26 (i.e., April to October 2025). When combining attempts supported by SSS and prescription medication, the prevalence of past-year quit attempts supported by SSS among all who smoked cigarettes was 3.1% and among those who tried to quit 7.8% in 2021/22 compared with 4.0% and 11.0% in 2025/26. Participant characteristics are presented in Table 1.

**Table 1:** Weighted characteristics of participants with complete data who smoked cigarettes in the past year (N_unweighted_=12,314).

|  | All smoking cigarettes<br>in past year | Those trying to<br>quit in past year |
| --- | --- | --- |
| Sample size, n | 13,398 | 5,076 |
| Age, median (IQR) | 40 (29, 55) | 35 (26, 51) |
| Women, % (95% CI) | 46.4 (45.5, 47.4) | 47.2 (45.6, 48.7) |
| Men, % (95% CI) | 52.2 (51.2, 53.1) | 51.3 (49.8, 52.9) |
| Non-binary, % (95% CI) | 1.4 (1.2, 1.6) | 1.7 (1.1, 1.8) |
| Less advantaged socioeconomic position, % (95% CI) | 58.0 (57.1, 58.9) | 58.2 (56.7, 59.7) |
| SSS-supported quit attempt, % (95% CI) |  |  |
| - Financial year 2021/22<sup>1</sup> | 0.5 (0.1, 0.8) | 1.2 (0.3, 2.1) |
| - Financial year 2022/23 | 0.8 (0.5, 1.2) | 2.2 (1.3, 3.1) |
| - Financial year 2023/24 | 0.8 (0.5, 1.2) | 2.3 (1.4, 3.1) |
| - Financial year 2024/25 | 1.5 (1.0, 1.9) | 3.7 (1.4, 3.2) |
| - Financial year 2025/26<sup>2</sup> | 1.8 (1.1, 2.4) | 4.8 (3.1, 6.5) |
| SSS-supported quit attempt (incl. meds), % (95% CI) |  |  |
| - Financial year 2021/22<sup>1</sup> | 3.1 (2.1, 4.0) | 7.8 (5.4, 10.2) |
| - Financial year 2022/23 | 2.2 (1.7, 2.8) | 5.8 (4.4, 7.2) |
| - Financial year 2023/24 | 2.5 (1.9, 3.1) | 6.9 (5.3, 8.5) |
| - Financial year 2024/25 | 3.5 (2.8, 4.2) | 8.9 (7.2, 10.7) |
| - Financial year 2025/26<sup>2</sup> | 4.0 (3.1, 5.0) | 11.0 (8.5, 13.6) |

Abbreviations: CI, confidence interval; IQR, interquartile range; SSS, Stop Smoking Service; meds, prescription medication to stop smoking. <sup>1</sup>October 2021 to March 2022. <sup>2</sup>April to October 2025. The financial year starts in April and ends in March the following year.

### Change in quit attempts supported by SSS

For the primary model, anticipating the intervention effect to start in April 2024 (i.e., start of financial year 2024) and using past-year quit attempts supported by SSS as the outcome, the step change was not statistically significant (Table 2, details in supplementary Table S2). When modelling an early intervention effect in February 2024, the step change was not statistically significant, but the β coefficient was larger, suggesting that an early effect might have been more likely (details in supplementary Table S3). This was further indicated by a complete attenuation of the step change when modelling it in July 2024 (see supplementary Table S4). There was no indication of a change in the trend following the implementation in April 2024 (see supplementary Table S5).

**Table 2:** Results for the association of increased SSS funding and prevalence of quit attempts using supported by SSS among adults who smoked cigarettes and tried to quit in the past year in England.

| Outcome | Step change | Unadjusted |  | Adjusted<sup>1</sup> |  |
| --- | --- | --- | --- | --- | --- |
| | | $\beta$ (95% CI) | OR (95% CI) | $\beta$ (95% CI) | OR (95% CI) |
| Past-year quit attempt supported by SSS | April 2024 | 0.142<br>(□0.536, 0.820) | 1.15 (0.59, 2.27) | 0.127<br>(□0.659, 0.912) | 1.13 (0.52, 2.49) |
| Past-year quit attempt supported by SSS | Feb 2024 | 0.414<br>(□0.294, 1.122) | 1.51 (0.75, 3.07) | 0.852<br>(□0.381, 2.085) | 2.34 (0.68, 8.04) |
| Past-year quit attempt supported by SSS, incl. meds | April 2024 | 0.246<br>(□0.168, 0.644) | 1.28 (0.84, 1.93) | 0.188<br>(□0.291, 0.666) | 1.21 (0.75, 1.95) |
| Past-year quit attempt supported by SSS, long time-series (2006-2025) | April 2024 | 0.996<br>(0.715, 1.276) | 2.71 (2.04, 3.58) | 0.994<br>(0.706, 1.282) | 2.70 (2.03, 3.60) |

<sup>1</sup>Adjusted for above-inflation tax increases, Swap-to-Stop, age, gender, and social grade. Model for long time-series, only adjusted for age, gender, and social grade. For full model specifications, see supplementary Tables S2, S3, S6, S8. Abbreviations: CI, confidence interval; OR, odds ratio; SSS, Stop Smoking Services.

When extending the outcome to include prescription medication, the step change in April appeared more pronounced than when only assessing SSS-supported attempts (Table 2 and supplementary Table S6). The results for the model including all who smoked cigarettes in the past year were roughly comparable to those of the primary model (supplementary Table S7). When using the long time series starting in November 2006, the step change in April 2024 was statistically significant, indicating that since the additional funding came into effect, the odds of someone who was trying to quit smoking being supported by SSS was 2.7-times higher than without the additional funding (Table 2, Figure 1, and supplementary Table S8).

**Figure 1:**
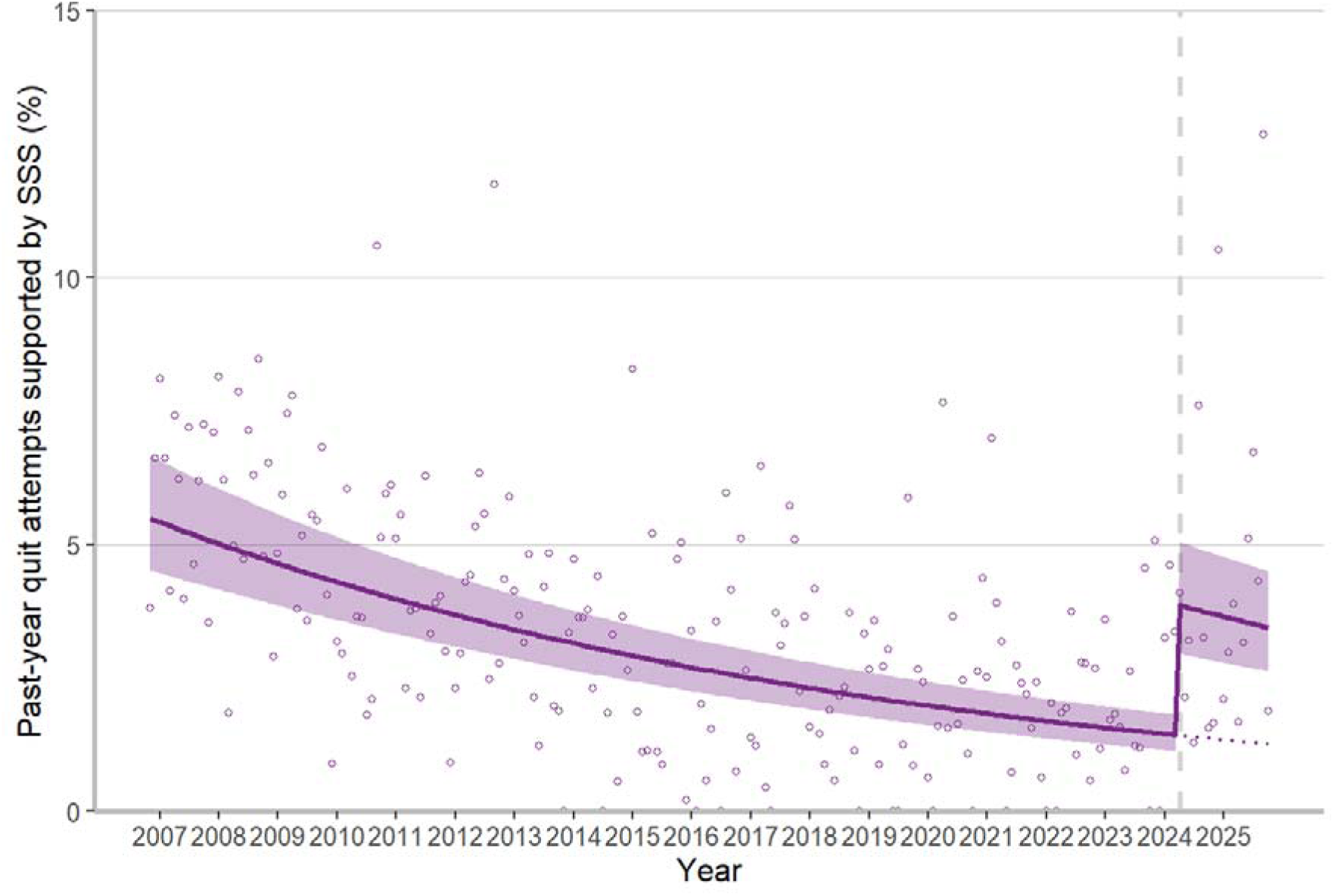
Modelled trends, before and after April 2024 (start of additional SSS funding in financial year 2024/25) in England, adjusted for seasonality, age, gender, and social grade. Dashed vertical line shows start of SSS funding. Dotted line indicates continued pre-intervention trend. Shaded areas indicate 95% confidence intervals and dots unmodelled estimates.

### Cost-effectiveness analysis

For the cost-effectiveness analysis, we used the primary model, which presented a lower effect size for the additional funding, as well as the long-term trends model, which indicated a higher effect size. The presented figures are based on estimated 6,575,861 people smoking cigarettes in England in 2024 and a permanent success rate of 2.5%.

Using the primary model, the overall ICER across the entire adult population was uncertain ranging from cost-effective to ineffective (£104,126; 95% CI: £939,398 to £8,293; Table 3). This value is based on an estimated effect size of 0.4% overall, with the highest estimate for the 45–54-year-old age group and the lowest for the 18–34-year-old age group. These were also the age groups with the highest and lowest cost-effectiveness, respectively. In terms of public health impact, the additional SSS funding resulted in an estimated 596 people who smoked cigarettes stopping permanently or 672 attributable DLYs gained.

**Table 3:** Calculations for incremental cost-effectiveness ratios and discounted life years across age bands for additional SSS funding.

|  | Time series | Age bands (years) |  |  |  |  | Overall<sup>1</sup> |
| --- | --- | --- | --- | --- | --- | --- | --- |
|  |  | 18-34 | 35-44 | 45-54 | 55-64 | ≥65 |  |
| Cigarette smoking, % (95% CI) | – | 18.8 (17.6, 19.9) | 15.9 (14.4, 17.4) | 14.3 (12.9, 15.7) | 13.1 (11.8, 14.4) | 7.8 (7.0, 8.6) | 14.2 (13.6, 14.7) |
| Additional SSS funding effect size, % (95% CI) | 2021-2025 | 0.30 (-0.86, 2.86) | 0.49 (-1.44, 4.58) | 0.96 (-2.74, 8.60) | 0.78 (-2.29, 6.98) | 0.82 (-2.41, 7.45) | 0.36 (-1.49, 3.93) |
|  | 2006-2025 | 1.88 (1.20, 2.76) | 2.80 (1.77, 4.12) | 3.38 (2.14, 4.97) | 4.40 (2.79, 6.40) | 4.15 (2.61, 6.12) | 2.72 (1.76, 3.92) |
| ICER, £ (95% CI) | 2021-2025 | 134,474 (1,410,483; 11,999) | 54,866 (612,706; 5,110) | 28,040 (303,876; 2,824) | 47,089 (501,844; 4,668) | – | 104,126 (939,398; 8,293) |
|  | 2006-2025 | 20,939 (32,858; 14,257) | 9,588 (15,143; 6,506) | 7,995 (12,636; 5,438) | 8,343 (13,138; 5,730) | – | 13,857 (21,393; 9,620) |
| Permanently stopping due to additional SSS funding, n (95% CI) | 2021-2025 | 179 (0; 1,651) | 154 (0; 1,365) | 248 (0; 2,054) | 189 (0; 1,575) | 176 (0; 1,496) | 596 (0; 6,498) |
|  | 2006-2025 | 1,134 (688; 1,741) | 881 (523; 1,372) | 869 (513; 1,354) | 1,068 (634; 1,653) | 889 (521; 1,391) | 4,477 (2,891; 6,449) |
| Attributable DLY gained, years (95% CI) | 2021-2025 | 194 (0; 1,790) | 245 (0; 2,167) | 391 (0; 3,239) | 220 (0; 1,828) | 0 | 672 (0; 7,332) |
|  | 2006-2025 | 1,230 (745; 1,888) | 1,400 (831; 2,179) | 1,371 (809; 2,136) | 1,240 (737; 1,919) | 0 | 5,052 (3,261; 7,276) |

<sup>1</sup> These estimates were derived separately and therefore may not exactly match the aggregated age-band estimates. Abbreviations: DLY, discounted life years; ICER, incremental cost-effectiveness ratio; SSS, Stop Smoking Service. Details of the calculation are provided in supplementary Tables S9-10.

Based on the long-term trends model, the overall ICER across the entire adult population was £13,857 with an estimated effect size of 2.7% (Table 3). This resulted in an estimated 4,477 people who smoked cigarettes stopping permanently or 5,052 attributable DLYs gained.

## DISCUSSION

### Summary of findings

This study assessed whether the additional funding allocated to SSS in England was associated with an increase in the number of SSS-supported quit attempts and explored the potential cost-effectiveness of this investment. In the primary model, the April 2024 step change, marking the start of the financial year in which the additional funding was implemented, was not statistically significant, which may have reflected an underpowered analysis. The direction of the effect suggested an uncertain increase in SSS-supported quit attempts in this primary analysis and in several sensitivity analyses.

As a result of these insensitive findings, the time series was extended back to 2006 to increase statistical power. This analysis showed a statistically significant step change in April 2024, suggesting that the odds of a quit attempt being supported by SSS increased 2.7-fold immediately following the introduction of the enhanced funding compared with the preceding period. This effect is more pronounced than in the other analyses, which may be explained by the longer time series better reflecting the long-term downward trend before the additional funding was provided.

The cost-effectiveness analysis showed considerable uncertainty, due to the imprecision of the effect size for the additional funding. In the primary model, the mean cost per additional DLY gained was £104,126 (95% CI: £939,398 to £8,293), reflecting wide bounds under lowlJ versus highlJeffectiveness assumptions. In the longlJterm trend model, the mean was £13,857 (95% CI: £21,393 to £9,620). These results should be interpreted alongside the National Institute for Health and Care Excellence’s (NICE) benchmark thresholds for costlJeffectiveness (from April 2026 £25,000–£35,000 per quality-adjusted life years gained), noting that our estimates are expressed per DLY gained rather than quality-adjusted life years (32).

### Comparison with previous research

Previous research showed the effectiveness of SSS for smoking cessation as well as their potential to reduce smoking-related health inequalities (8, 33). A modelling study estimated that around 15% of the decline in smoking prevalence in England between 2001 to 2016 may be attributable to SSS (34). However, public funding for SSS declined drastically from 2015-16 onward (35, 36). These cuts led some local authorities to discontinue specialist SSS entirely (35).

The re-investment in SSS funding in 2024-25 appears to have reversed parts of this trajectory. A report by Action on Smoking and Health shows that, following this funding boost, most local authorities increased both the number of advisers and the number of settings offering SSS support (37). Our findings align with this broader pattern: although the effect detected in our primary model was modest and not significant, the longer time series analysis indicated a significant post-intervention increase in the odds of a quit attempt being supported by SSS.

The policy environment during this period was also shaped by the launch of the Swap-to-Stop scheme in December 2023, which provides free vape starter kits with behavioural support, including through SSS. An evaluation of the scheme suggests it was associated with approximately 125,000 additional vape-supported quit attempts in its first year (38). Although we adjusted our primary analysis for Swap-to-Stop scheme, it is also possible that it had synergistic effects together with the additional SSS funding and jointly increased interest in SSS for smoking cessation.

Previous cost-effectiveness studies of SSS typically evaluated the service itself rather than incremental changes in funding and found SSS to be highly cost-effective (39, 40). Early estimates (2000–01) suggested a cost per life-year gained of £684, with even lower effect size assumptions yielding costs far below the NICE thresholds (39). Subsequent work also highlighted that specialist SSS models tend to yield the lowest cost per person quitting compared to other delivery settings such as general practice or dentistry (40).

Our study differs from this earlier literature in a crucial way: we evaluated the marginal costlJeffectiveness of additional SSS funding, not the inherent costlJeffectiveness of the service. This distinction helps explain why our estimates appear more uncertain and, in scenarios of low effectiveness, substantially higher. The longlJterm trend model, which may have better captured the underlying trajectory of SSS use, produced an estimate broadly consistent with established evidence showing that investment in smoking cessation is a highly costlJeffective use of public funds.

### Implications for policy

The UK Government is currently in the process of passing a new legislation, the Tobacco and Vapes Bill, which includes the smokefree generation policy. This policy will increase the legal age of sale of tobacco one year every year until the sale of tobacco products eventually becomes completely illegal. In the medium- to long-term this policy has the potential to significantly reduce smoking rates (41). However, legislative measures that prevent uptake must be paired with interventions that support smoking cessation. In this context, sustained and enhanced funding for SSS represents an evidence-based complementary approach. While our findings indicate some uncertainty, the longer-term modelling and the broader literature strongly suggest that increased investment in SSS is likely to be clinically and cost-effective in helping people quit smoking.

### Strengths and limitations

A key strength of this study is the use of a long-running, monthly, nationally representative survey. Among the limitations are that the primary analysis may not have been sufficiently powered to detect an effect. To address this, we conducted an unplanned sensitivity analysis extending the time series back to November 2006. In this extended model, we did not adjust for other tobacco control policies, as numerous external factors could have influenced the outcome. Therefore, the primary analysis was focussed on a more stable period starting in October 2021. Nonetheless, we believe that the modelling approach and longer time series likely smoothed out the influence of policy changes.

Another limitation is that the study only included 19 months of data following the introduction of the additional funding, meaning the post-intervention period was relatively short, and only provides short-term insights into changes. Future research should extend the follow-up time to capture longer-term impact. As the study used an interrupted time-series design with repeat cross-sectional data, the observed step change represents a temporal association rather than causal evidence that the funding led to increased quit attempt rates.

Moreover, we did not assess whether the additional funding translated into more people successfully quitting, although we explored its potential public health impact as part of the cost-effectiveness analysis. Additionally, our outcome measure captured quit attempts within the past year, meaning some quit attempts recorded during the intervention period may have occurred before the additional funding was introduced. In the protocol, we had planned a sensitivity analysis focussing solely on past-month quit attempts, but the number of SSS-supported quit attempts in a given month was too low to permit a robust analysis.

## CONCLUSIONS

Compared with the long-term trend, increased funding in 2024 appeared to lead to an increase in quit attempts supported by SSS in England at the population level. This result is somewhat uncertain because our primary pre-planned analyses assessing the impact relative to a more recent trend were insensitive. However, the long-term trend model starting in 2006 indicated almost three-times higher odds of using SSS in quit attempts compared to the period before the additional funding started. It also indicated that the additional funding was cost-effective against the NICE benchmarks, although these are measured in quality-adjusted life years while our estimates are per DLY and reflect incremental impact of additional funding rather than cost-effectiveness of SSS. Together with the already established evidence-base on the effectiveness of SSS and increased service throughput reported by SSS providers, these results support the policy case for sustained investment in SSS as part of England’s wider tobacco control strategy.

## ETHICAL APPROVAL

The University College London Ethics Committee granted ethical approval for the Smoking and Alcohol Toolkit Study (ID 0498/001). Participants provided informed consent prior to taking part in the study.

## DATA AVAILABILITY

The data that support the findings of this study will be made openly available upon publication of the article in Open Science Framework at https://osf.io/r59eu.

## AUTHOR CONTRIBUTION STATEMENT

VHB, LS, LB, SM, and JB all contributed to the conceptualisation and methodology of this work. VHB and JB were responsible for data curation. VHB conducted the formal analysis and writing of the original draft. JB validated the results of the analysis. LS, LB, SM, and JB acquired the funding for this study and reviewed and edited the manuscript.

During the preparation of this manuscript the authors used Microsoft Copilot to improve readability and language. After using this tool, the authors reviewed and edited the content as needed and take full responsibility for the content of the published article.

## DECALARATION OF COMPETING INTERESTS

LS has received honoraria for talks, unrestricted research grants and travel expenses to attend meetings and workshops from manufactures of smoking cessation medications (Pfizer; J&J) and has acted as paid reviewer for grant awarding bodies and as a paid consultant for health care companies. All authors declare no financial links with the tobacco or vaping industry or their representatives.

