## Supplementary Material for "Impact of increased funding for Stop Smoking Services in England on quit attempts: population-based study 2021-2025"

### Methods

#### Missing values

If the exact age was missing (n=218), but participants stated the age band they belonged to, we imputed their age with the median age based on those in their age band that provided the exact age.

**Table S1:** Variables used for the analysis and the number (%) of missing values for each of them (people who smoked in past year in England, $N_{unweighted}$=12,882).

| Variable | Missing values n (%) |
|---|---|
| Exact age | 218 (1.7) |
| Age banded | 3 (0.0) |
| Gender | 76 (0.6) |
| Social grade | 0 (0) |
| Quit attempt in past year | 496 (3.9) |
| Quit attempt in past month | 0 (0) |
| Using SSS in past-year quit attempt | 0 (0) |
| Using SSS including medication in past-year quit attempt | 0 (0) |

#### R Packages

The following R packages and their dependencies were used in the analysis: tidyverse [1]; survey [2]; tibbletime [3]; ggplot2 [4]; zoo [5]; splines [6]; splines2 [7]; mgcv [8]; haven [9].

#### Quit attempts supported by SSS or prescription medication, 2007-2025

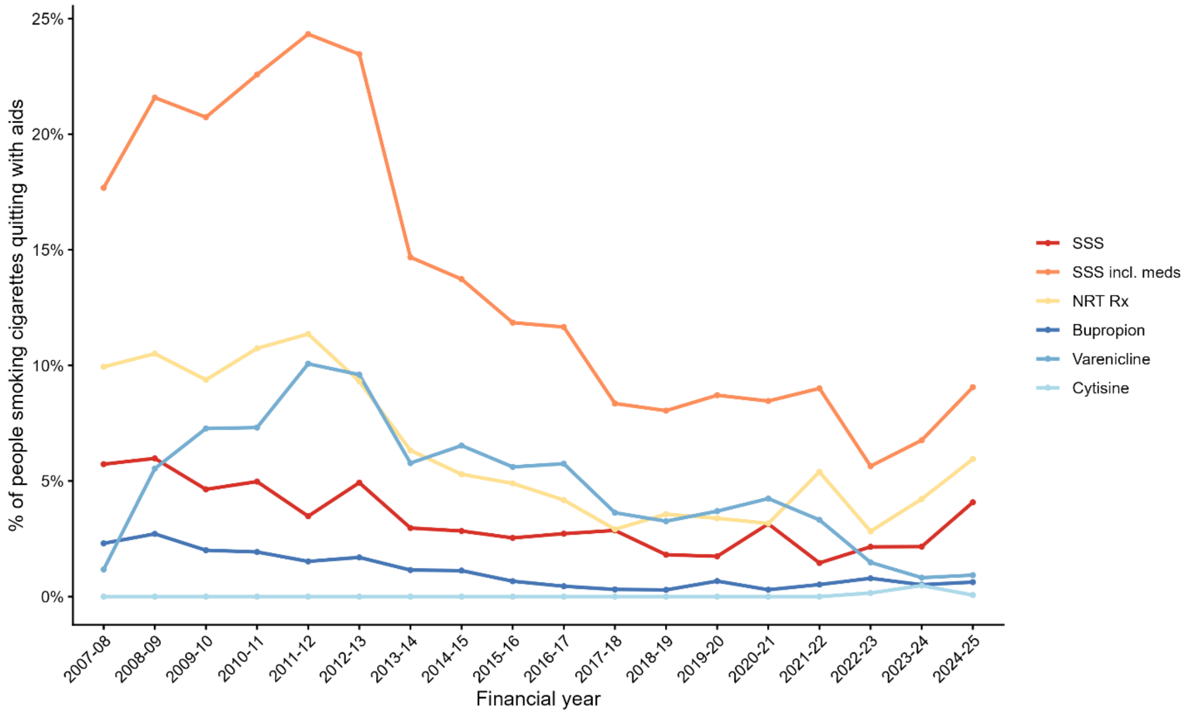

**Figure S1:** Quit attempts supported by SSS or prescription medication among people who smoked cigarettes in the past year. Weighted percentage per financial year. Abbreviations: NRT Rx, nicotine replacement products on prescription; SSS, Stop Smoking Service; SSS incl. meds, Stop Smoking Service including prescription medication.

#### Primary model: step change in April

Model to assess impact of increased funding for Stop Smoking Services (SSS) on past-year quit attempts supported by SSS among people who smoked cigarettes and tried to quit in the past year.

Model specifications:

- Generalised Additive Model (GAM) using gam-function of mgcv package [8]
- Family: quasibinomial; link function: logit; smooth term: cyclic cubic regression spline
- Method: restricted maximum likelihood; optimizer: outer newton
- Formula (only adjusted for seasonality): past-year quit attempt supported by SSS ~ step + time + s(seasonality, k = 12)

**Table S2**: Parameter estimates for GAM with prevalence of past-year quit attempt supported by SSS as outcome and step change in April 2024.

| **Variables** | **Unadjusted[1] model** | | | **Adjusted[2] model** | | |
|---|---|---|---|---|---|---|
| | β | p-value | OR (95% CI) | β | p-value | OR (95% CI) |
| Intercept | -4.29 | <0.001 | 0.01 (0.01, 0.02) | -6.29 | <0.001 | 0.00 (0.00, 0.00) |
| Time trend | 0.02 | 0.059 | 1.02 (1.00, 1.05) | 0.02 | 0.111 | 1.02 (0.99, 1.06) |
| Step change in April | 0.14 | 0.681 | 1.15 (0.59, 2.27) | 0.13 | 0.752 | 1.13 (0.52, 2.49) |

[1]Adjusted for seasonality. [2]Adjusted for seasonality, tobacco tax increases, Swap to Stop, age, gender, social grade. Abbreviation: CI, confidence interval; OR, odds ratio.

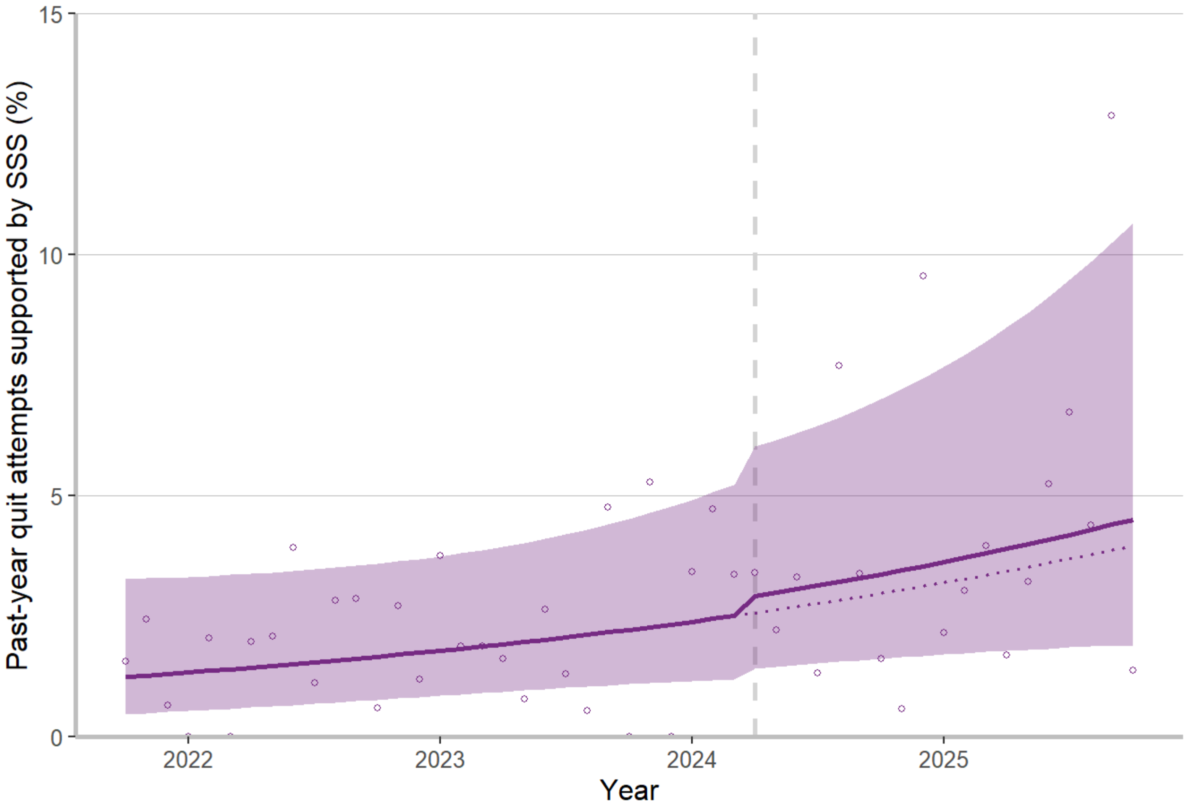

**Figure S2:** Modelled trends before and after April 2024 (start of additional SSS funding) in England, adjusted for seasonality, tobacco tax increases, Swap to Stop, age, gender, and social grade. Dashed vertical line shows start of SSS funding (intervention). Dotted line indicates continued pre-intervention trend. Shaded areas indicate 95% confidence intervals and dots unmodelled estimates.

#### Sensitivity analysis 1: step change in February

Model specifications:

- GAM using gam-function of mgcv package [8]
- Family: quasibinomial; link function: logit; smooth term: cyclic cubic regression spline
- Method: restricted maximum likelihood; optimizer: outer newton
- Formula (only adjusted for seasonality): past-year quit attempt supported by SSS ~ step + time + s(seasonality, k = 12)

**Table S3**: Parameter estimates for GAM with prevalence of past-year quit attempt supported by SSS as outcome and step change in February 2024.

| **Variables** | **Unadjusted[1] model** | | | **Adjusted[2] model** | | |
|---|---|---|---|---|---|---|
| | β | p-value | OR (95% CI) | β | p-value | OR (95% CI) |
| Intercept | -4.20 | <0.001 | 0.01 (0.01, 0.02) | -6.02 | <0.001 | 0.00 (0.00, 0.01) |
| Time trend | 0.02 | 0.240 | 1.02 (0.99, 1.04) | 0.02 | 0.102 | 1.02 (1.00, 1.05) |
| Step change in February | 0.41 | 0.252 | 1.51 (0.75, 3.07) | 0.89 | 0.154 | 2.43 (0.72, 8.24) |

[1]Adjusted for seasonality. [2]Adjusted for seasonality, tobacco tax increases, Swap to Stop, age, gender, social grade. Abbreviation: CI, confidence interval; OR, odds ratio.

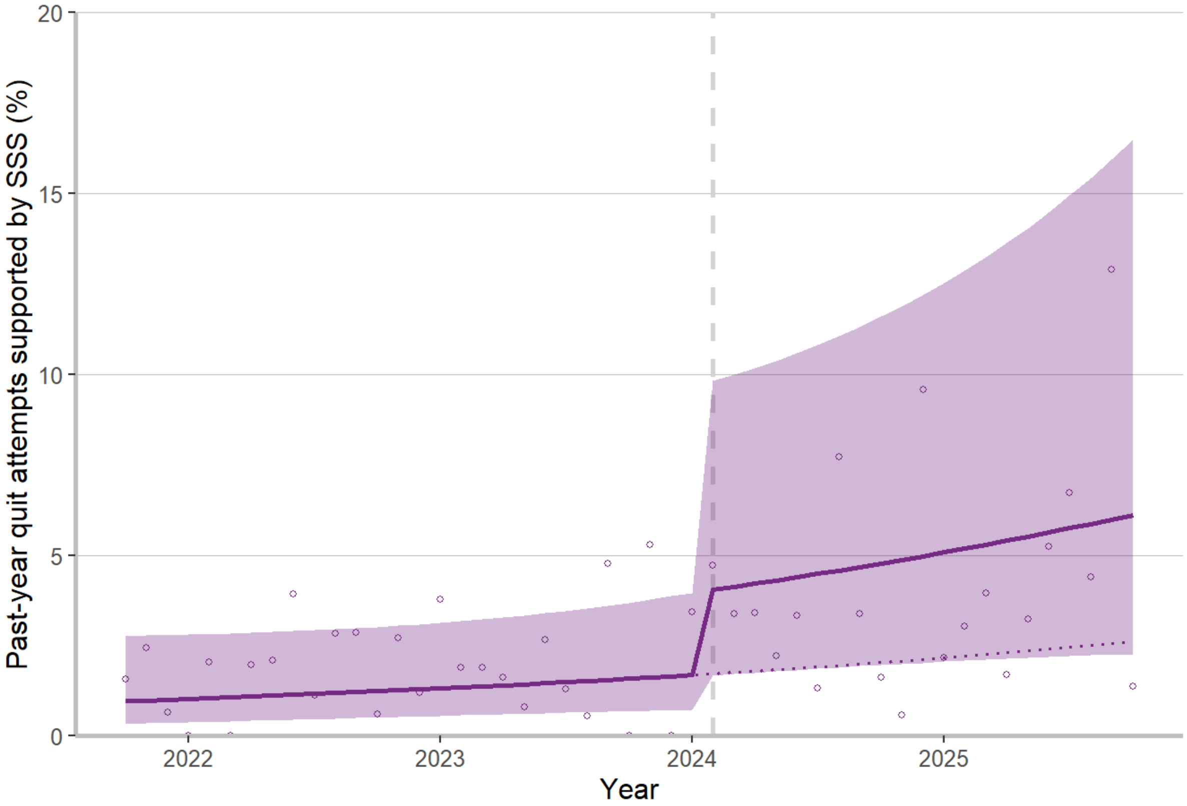

**Figure S3:** Modelled trends before and after February 2024 (start of additional SSS funding) in England, adjusted for seasonality, tobacco tax increases, Swap to Stop, age, gender, and social grade. Dashed vertical line shows start of SSS funding (intervention). Dotted line indicates continued pre-intervention trend. Shaded areas indicate 95% confidence intervals and dots unmodelled estimates.

#### Sensitivity analysis 2: step change in July

Model specifications:

- GAM using gam-function of mgcv package [8]
- Family: quasibinomial; link function: logit; smooth term: cyclic cubic regression spline
- Method: restricted maximum likelihood; optimizer: outer newton
- Formula (only adjusted for seasonality): past-year quit attempt supported by SSS ~ step + time + s(seasonality, k = 12)

**Table S4**: Parameter estimates for GAM with prevalence of past-year quit attempt supported by SSS as outcome and step change in July 2024.

| **Variables** | **Unadjusted[1] model** | | | **Adjusted[2] model** | | |
|---|---|---|---|---|---|---|
| | β | p-value | OR (95% CI) | β | p-value | OR (95% CI) |
| Intercept | -4.34 | <0.001 | 0.01 (0.01, 0.02) | -6.29 | <0.001 | 0.00 (0.00, 0.01) |
| Time trend | 0.03 | 0.019 | 1.03 (1.01, 1.05) | 0.02 | 0.136 | 1.03 (0.99, 1.06) |
| Step change in July | 0.01 | 0.979 | 1.01 (0.53, 1.90) | 0.04 | 0.898 | 1.04 (0.55, 1.98) |

[1]Adjusted for seasonality. [2]Adjusted for seasonality, tobacco tax increases, Swap to Stop, age, gender, social grade. Abbreviation: CI, confidence interval; OR, odds ratio.

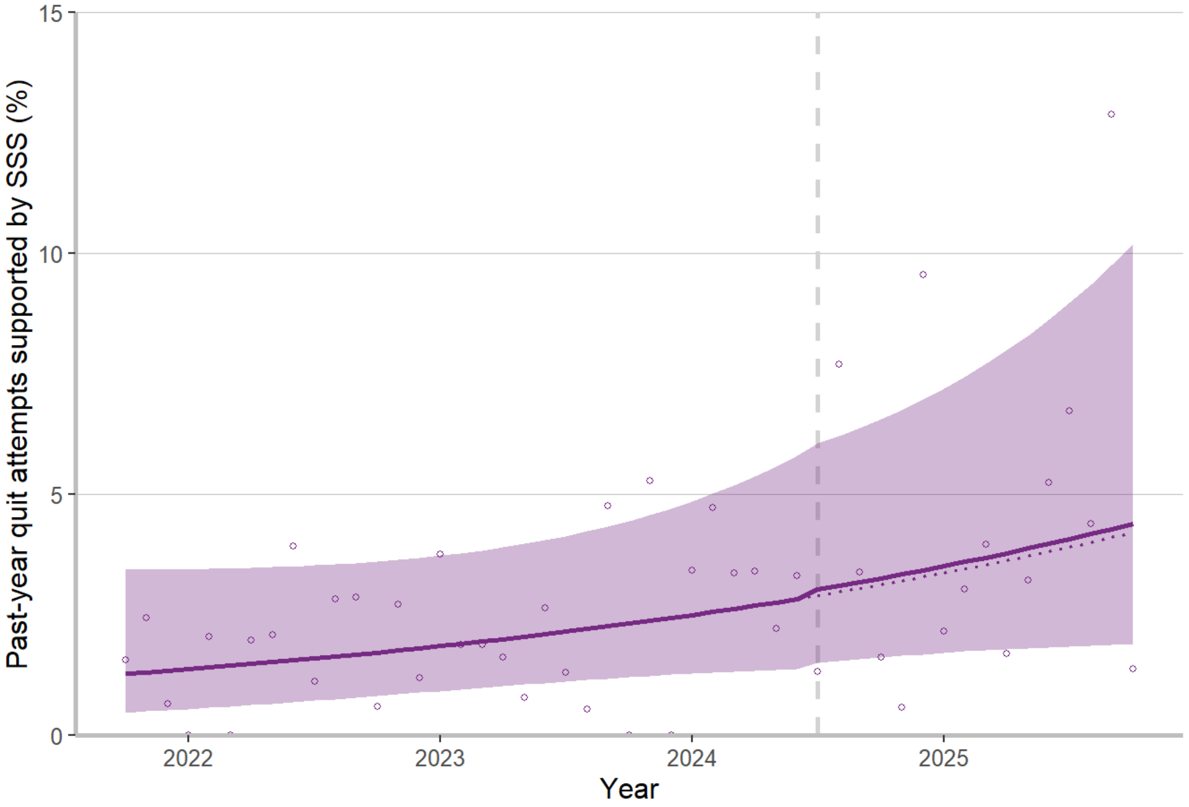

**Figure S4:** Modelled trends before and after July 2024 (start of additional SSS funding) in England, adjusted for seasonality, tobacco tax increases, Swap to Stop, age, gender, and social grade. Dashed vertical line shows start of SSS funding (intervention). Dotted line indicates continued pre-intervention trend. Shaded areas indicate 95% confidence intervals and dots unmodelled estimates.

#### Sensitivity analysis 3: trend change

Model specifications:

- GAM using gam-function of mgcv package [8]
- Family: quasibinomial; link function: logit; smooth term: cyclic cubic regression spline
- Method: restricted maximum likelihood; optimizer: outer newton
- Formula (only adjusted for seasonality): past-year quit attempt supported by SSS ~ step*time + s(seasonality, k = 12)

**Table S5**: Parameter estimates for GAM with prevalence of past-year quit attempt supported by SSS as outcome and step change in April 2024.

| Variables | Unadjusted[1] model | | | Adjusted[2] model | | |
|---|---|---|---|---|---|---|
| | β | p-value | OR (95% CI) | β | p-value | OR (95% CI) |
| Intercept | -4.27 | <0.001 | 0.01 (0.01, 0.02) | -6.16 | <0.001 | 0.00 (0.00, 0.01) |
| Time trend | 0.02 | 0.143 | 1.02 (0.99, 1.05) | 0.01 | 0.485 | 1.01 (0.97, 1.06) |
| Step change in April | 0.04 | 0.964 | 1.04 (0.17, 6.23) | -0.56 | 0.622 | 0.57 (0.06, 5.30) |
| Change in trend | 0.00 | 0.905 | 1.00 (0.95, 1.06) | 0.02 | 0.518 | 1.02 (0.96, 1.08) |

[1]Adjusted for seasonality. [2]Adjusted for seasonality, tobacco tax increases, Swap to Stop, age, gender, social grade. Abbreviation: CI, confidence interval; OR, odds ratio.

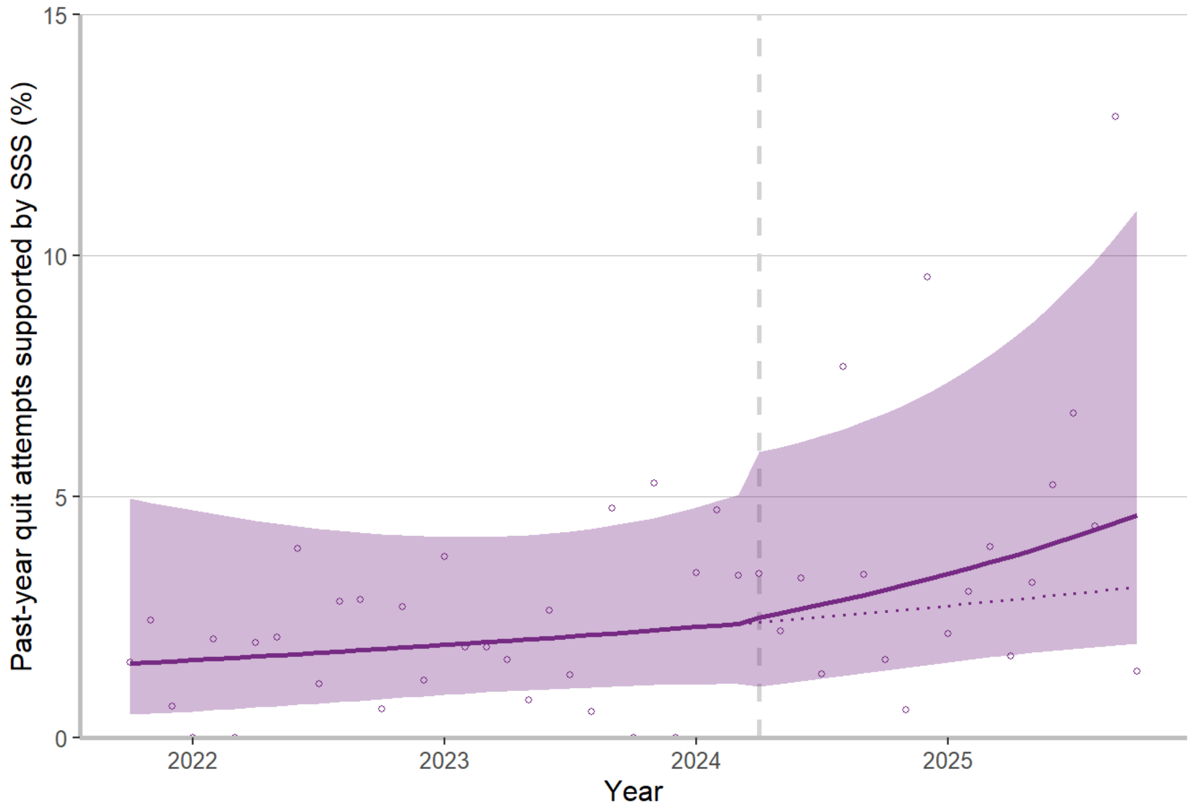

**Figure S5:** Modelled trends before and after April 2024 (start of additional SSS funding) in England, assuming a change in trend, adjusted for seasonality, tobacco tax increases, Swap to Stop, age, gender, and social grade. Dashed vertical line shows start of SSS funding (intervention). Dotted line indicates continued pre-intervention trend. Shaded areas indicate 95% confidence intervals and dots unmodelled estimates.

#### Sensitivity analysis 4: quit attempts supported by SSS incl. medication

Model specifications:

- GAM using gam-function of mgcv package [8]
- Family: quasibinomial; link function: logit; smooth term: cyclic cubic regression spline
- Method: restricted maximum likelihood; optimizer: outer newton
- Formula (only adjusted for seasonality): past-year quit attempt supported by SSS including medication ~ step + time + s(seasonality, k = 12)

**Table S6**: Parameter estimates for GAM with prevalence of past-year quit attempt supported by SSS, including prescription stop smoking medication, as outcome and step change in April 2024.

| **Variables** | **Unadjusted[1] model** | | | **Adjusted[2] model** | | |
|---|---|---|---|---|---|---|
| | β | p-value | OR (95% CI) | β | p-value | OR (95% CI) |
| Intercept | -2.74 | <0.001 | 0.06 (0.05, 0.08) | -4.73 | <0.001 | 0.01 (0.01, 0.01) |
| Time trend | 0.01 | 0.377 | 1.01 (0.99, 1.02) | 0.01 | 0.356 | 1.01 (0.99, 1.03) |
| Step change in April | 0.25 | 0.245 | 1.28 (0.84, 1.93) | 0.19 | 0.443 | 1.21 (0.75, 1.95) |

[1]Adjusted for seasonality. [2]Adjusted for seasonality, tobacco tax increases, Swap to Stop, age, gender, social grade. Abbreviation: CI, confidence interval; OR, odds ratio.

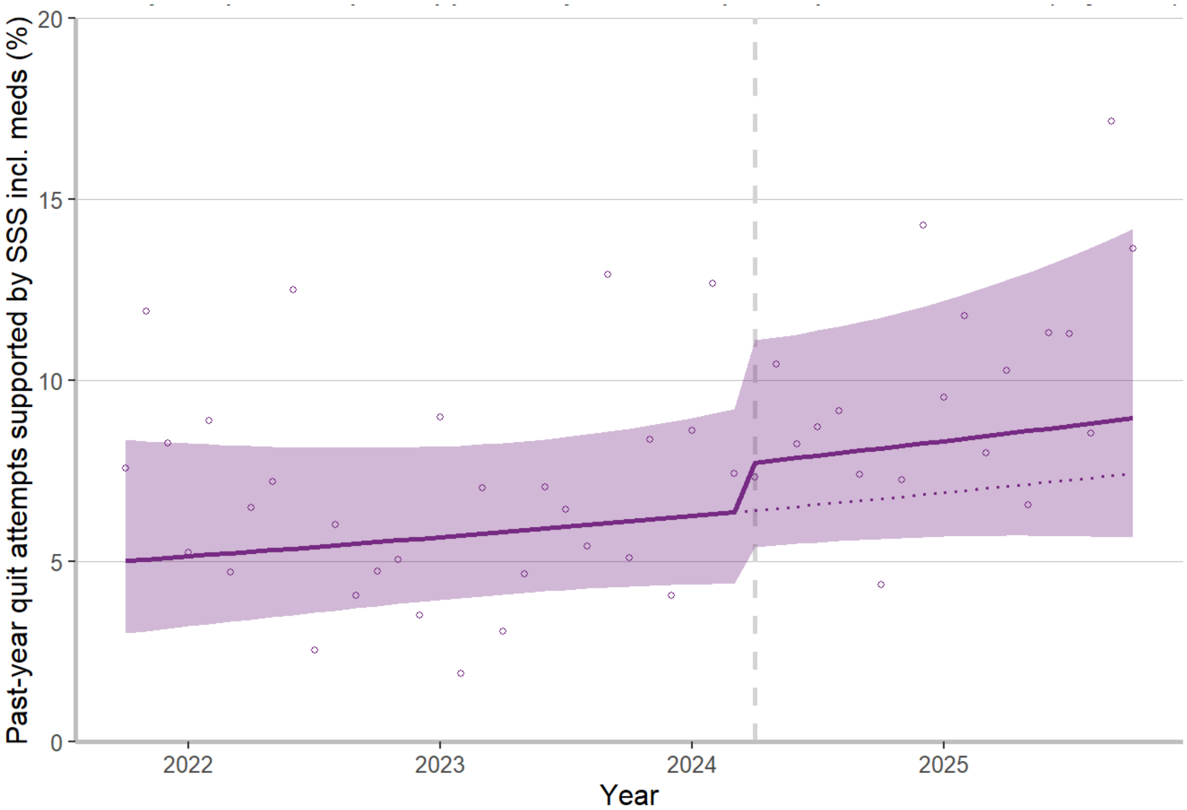

**Figure S6:** Modelled trends before and after February 2024 (start of additional SSS funding) in England, adjusted for seasonality, tobacco tax increases, Swap to Stop, age, gender, and social grade. Outcome includes use of prescription stop smoking medication. Dashed vertical line shows start of SSS funding (intervention). Dotted line indicates continued pre-intervention trend. Shaded areas indicate 95% confidence intervals and dots unmodelled estimates.

#### Sensitivity analysis 5: all who smoked in past year

Model specifications:

- GAM using gam-function of mgcv package [8]
- Family: quasibinomial; link function: logit; smooth term: cyclic cubic regression spline
- Method: restricted maximum likelihood; optimizer: outer newton
- Formula (only adjusted for seasonality): past-year quit attempt supported by SSS ~ step + time + s(seasonality, k = 12)

**Table S7**: Parameter estimates for GAM with prevalence of past-year quit attempt supported by SSS among all who smoked cigarettes in past year as outcome and step change in April 2024.

| Variables | Unadjusted[1] model | | | Adjusted[2] model | | |
|---|---|---|---|---|---|---|
| | β | p-value | OR (95% CI) | β | p-value | OR (95% CI) |
| Intercept | -5.23 | <0.001 | 0.01 (0.00, 0.01) | -6.83 | <0.001 | 0.00 (0.00, 0.00) |
| Time trend | 0.02 | 0.127 | 1.02 (0.99, 1.04) | 0.02 | 0.249 | 1.02 (0.99, 1.05) |
| Step change in April | 0.26 | 0.444 | 1.30 (0.66, 2.54) | 0.26 | 0.511 | 1.30 (0.60, 2.82) |

[1]Adjusted for seasonality. [2]Adjusted for seasonality, tobacco tax increases, Swap to Stop, age, gender, social grade. Abbreviation: CI, confidence interval; OR, odds ratio.

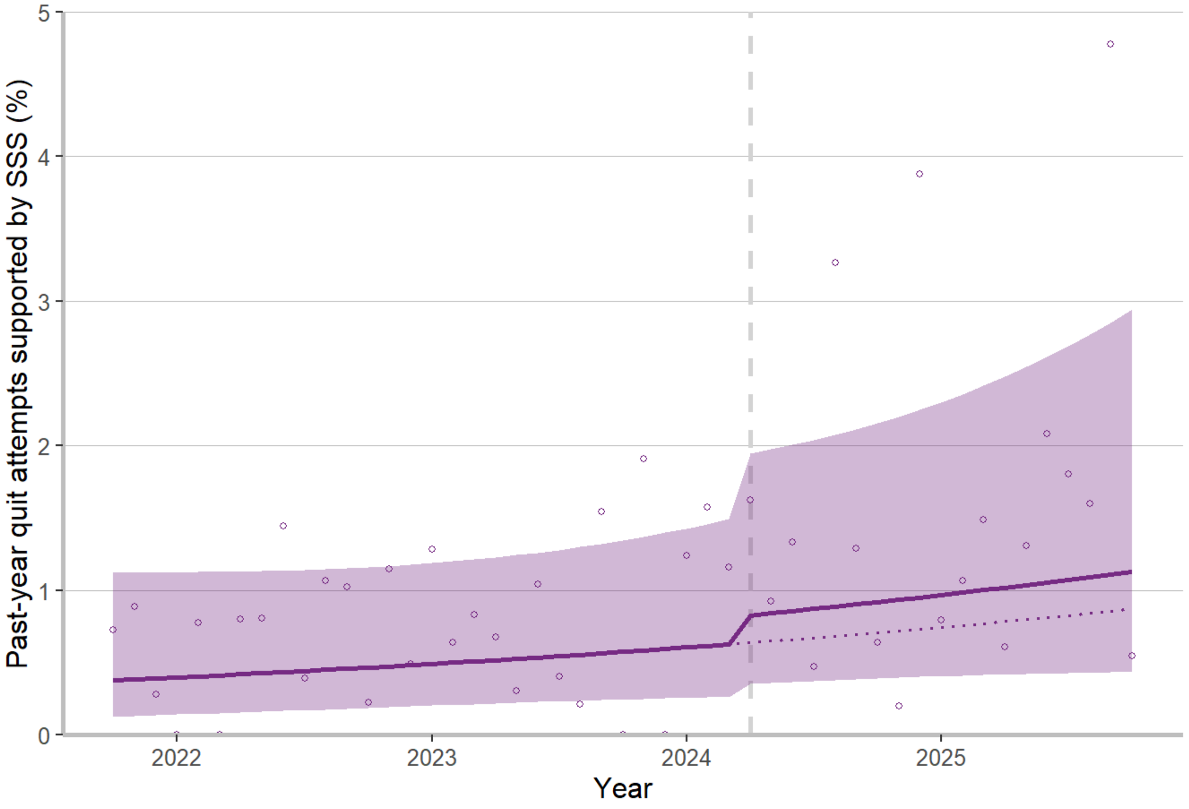

**Figure S7:** Modelled trends among all who cigarettes smoked in the past year, before and after April 2024 (start of additional SSS funding) in England, adjusted for seasonality, tobacco tax increases, Swap to Stop, age, gender, and social grade. Dashed vertical line shows start of SSS funding (intervention). Dotted line indicates continued pre-intervention trend. Shaded areas indicate 95% confidence intervals and dots unmodelled estimates.

#### Sensitivity analysis 6: Time series starting in November 2006

Model specifications:

- GAM using gam-function of mgcv package [8]
- Family: quasibinomial; link function: logit; smooth term: cyclic cubic regression spline
- Method: restricted maximum likelihood; optimizer: outer newton
- Formula (only adjusted for seasonality): past-year quit attempt supported by SSS ~ step + time + s(seasonality, k = 12)

**Table S8**: Parameter estimates for GAM with prevalence of past-year quit attempt supported by SSS as outcome and step change in April 2024, using a long series starting in November 2006.

| **Variables** | **Unadjusted[1] model** | | | **Adjusted[2] model** | | |
|---|---|---|---|---|---|---|
| | β | p-value | OR (95% CI) | β | p-value | OR (95% CI) |
| Intercept | -2.74 | <0.001 | 0.06 (0.06, 0.07) | -3.82 | <0.001 | 0.02 (0.02, 0.03) |
| Time trend | -0.01 | <0.001 | 0.99 (0.99, 0.99) | -0.01 | <0.001 | 0.99 (0.99, 0.99) |
| Step change in April | 1.00 | <0.001 | 2.71 (2.04, 3.58) | 0.99 | <0.001 | 2.70 (2.03, 3.60) |

[1]Adjusted for seasonality. [2]Adjusted for seasonality, age, gender, social grade. Abbreviation: CI, confidence interval; OR, odds ratio.

#### Cost-effectiveness analysis

**Table S9:** Calculations for incremental cost-effectiveness ratios and discounted life years across age bands for additional SSS funding, using primary analysis (time series from 2021-2025 – pessimistic effect size).

| | Age bands (years) | | | | | |
|---|---|---|---|---|---|---|
| | 18-34 | 35-44 | 45-54 | 55-64 | ≥65 | Overall[6] |
| **A.** % (95% CI) SSS effect size | 0.30 (-0.86, 2.86) | 0.49 (-1.44, 4.58) | 0.96 (-2.74, 8.60) | 0.78 (-2.29, 6.98) | 0.82 (-2.41, 7.45) | 0.36 (-1.49, 3.93) |
| **B.** % quitting permanently | 2.5 | 2.5 | 2.5 | 2.5 | 2.5 | 2.5 |
| **C.** DLY gained attributable to SSS/ person stopping permanently[1] | 1.084 | 1.588 | 1.577 | 1.161 | 0 | 1.128 |
| **D.** % (95% CI) smoking cigarettes in England (2024) | 18.8 (17.6, 19.9) | 15.9 (14.4, 17.4) | 14.3 (12.9, 15.7) | 13.1 (11.8, 14.4) | 7.8 (7.0, 8.6) | 14.2 (13.6, 14.7) |
| **E.** Population England (mid-2024)[2] | 12,894,571 | 7,928,738 | 7,208,565 | 7,424,119 | 10,981,092 | 46,437,085 |
| **F.** (D*E) = number (95% CI) smoking cigarettes | 2,418,604 (2,264,810; 2,572,399) | 1,260,569 (1,142,756; 1,378,383) | 1,029,543 (929,653; 1,129,433) | 972,192 (875,524; 1,068,860) | 856,656 (764,584; 948,728) | 6,575,861 (6,317,063; 6,834,660) |
| **G.** £ cost per person smoking[3] | 10.64 | 10.64 | 10.64 | 10.64 | 10.64 | 10.64 |
| **H.** (A*B) = % (95% CI) stopping permanently due to SSS | 0.01 (-0.02, 0.07) | 0.01 (-0.04, 0.11) | 0.02 (-0.07, 0.22) | 0.02 (-0.06, 0.17) | 0.02 (-0.06, 0.19) | 0.01 (-0.04, 0.10) |
| **I.** (C*H)/100 = $10^{-3}$ mean attributable DLY gained/person smoking[3] | 0.08 (0, 0.78) | 0.19 (0, 1.82) | 0.38 (0, 3.39) | 0.23 (0, 2.09) | 0 | 0.10 (0, 1.11) |
| **J.** (I/G) £ ICER (95% CI)[4] | 134,474 (1,410,483; 11,999) | 54,866 (612,706; 5,110) | 28,040 (303,876; 2,824) | 47,089 (501,844; 4,668) | – | 104,126 (939,398; 8,293) |

| | Age bands (years) | | | | | |
|---|---|---|---|---|---|---|
| | 18-34 | 35-44 | 45-54 | 55-64 | ≥65 | Overall[6] |
| **K.** (F*H) = number (95% CI) stopping permanently due to SSS | 179 (0; 1,651) | 154 (0; 1,365) | 248 (0; 2,054) | 189 (0; 1,575) | 176 (0; 1,496) | 596 (0; 6,498) |
| **L.** (C*K) = attributable DLY gained | 194 (0; 1,790) | 245 (0; 2,167) | 391 (0; 3,239) | 220 (0; 1,828) | 0 | 672 (0; 7,332) |

[1] Taken from [10]. [2] Using mid-2024 population estimates from Office for National Statistics [11]. [3] The Government estimated the cost per person as £12.39 based on 5.6 million people smoking in England and a total cost of £70 million [12]. However, this is based on Annual Population Survey smoking rates which may be an underestimation as it may not capture all people smoking non-daily [13]. Therefore, we used estimates from our survey. [4] Negative value for lower bound of 95% CI set to 0. [5] Non-infinite values were removed for calculation of lower bound of 95% CI – an effect size of ≥0 suggests additional funding did not result in more people quitting. [6] These estimates were derived separately and therefore may not exactly match the aggregated age-band estimates. Abbreviations: CI, confidence interval; DLY, discounted life years; ICER, incremental cost-effectiveness ratio; SSS, Stop Smoking Service.

**Table S10:** Calculations for incremental cost-effectiveness ratios and discounted life years across age bands for additional SSS funding, using primary analysis (time series from 2006-2025 – optimistic effect size).

| | Age bands (years) | | | | | |
|---|---|---|---|---|---|---|
| | 18-34 | 35-44 | 45-54 | 55-64 | ≥65 | Overall[5] |
| **A.** % (95% CI) SSS effect size | 1.88 (1.20, 2.76) | 2.80 (1.77, 4.12) | 3.38 (2.14, 4.97) | 4.40 (2.79, 6.40) | 4.15 (2.61, 6.12) | 2.72 (1.76, 3.92) |
| **B.** % quitting permanently | 2.5 | 2.5 | 2.5 | 2.5 | 2.5 | 2.5 |
| **C.** DLY gained attributable to SSS/ person stopping permanently[1] | 1.084 | 1.588 | 1.577 | 1.161 | 0 | 1.128 |
| **D.** % (95% CI) smoking cigarettes in England (2024) | 18.8 (17.6, 19.9) | 15.9 (14.4, 17.4) | 14.3 (12.9, 15.7) | 13.1 (11.8, 14.4) | 7.8 (7.0, 8.6) | 14.2 (13.6, 14.7) |
| **E.** Population England (mid-2024)[2] | 12,894,571 | 7,928,738 | 7,208,565 | 7,424,119 | 10,981,092 | 46,437,085 |

| | Age bands (years) | | | | | |
|---|---|---|---|---|---|---|
| | 18-34 | 35-44 | 45-54 | 55-64 | ≥65 | Overall[5] |
| **F.** (D*E) = number (95% CI) smoking cigarettes | 2,418,604 (2,264,810; 2,572,399) | 1,260,569 (1,142,756; 1,378,383) | 1,029,543 (929,653; 1,129,433) | 972,192 (875,524; 1,068,860) | 856,656 (764,584; 948,728) | 6,575,861 (6,317,063; 6,834,660) |
| **G.** £ cost per person smoking[3] | 10.64 | 10.64 | 10.64 | 10.64 | 10.64 | 10.64 |
| **H.** (A*B) = % (95% CI) stopping permanently due to SSS | 0.05 (0.03, 0.07) | 0.07 (0.04, 0.10) | 0.08 (0.05, 0.12) | 0.11 (0.07, 0.16) | 0.10 (0.07, 0.15) | 0.07 (0.04, 0.10) |
| **I.** (C*H)/100 = $10^{-3}$ mean attributable DLY gained/person smoking[3] | 0.51 (0.32, 0.75) | 1.11 (0.70, 1.64) | 1.33 (0.84, 1.96) | 1.28 (0.81, 1.86) | 0 | 0.77 (0.50, 1.11) |
| **J.** (I/G) £ ICER (95% CI)[4] | 20,939 (32,858; 14,257) | 9,588 (15,143; 6,506) | 7,995 (12,636; 5,438) | 8,343 (13,138; 5,730) | – | 13,857 (21,393; 9,620) |
| **K.** (F*H) = number (95% CI) stopping permanently due to SSS | 1,134 (688; 1,741) | 881 (523; 1,372) | 869 (513; 1,354) | 1,068 (634; 1,653) | 889 (521; 1,391) | 4,477 (2,891; 6,449) |
| **L.** (C*K) = attributable DLY gained | 1,230 (745; 1,888) | 1,400 (831; 2,179) | 1,371 (809; 2,136) | 1,240 (737; 1,919) | 0 | 5,052 (3,261; 7,276) |

[1] Taken from [10]. [2] Using mid-2024 population estimates from Office for National Statistics [11]. [3] The Government estimated the cost per person as £12.39 based on 5.6 million people smoking in England and a total cost of £70 million [12]. However, this is based on Annual Population Survey smoking rates which may be an underestimation as it may not capture all people smoking non-daily [13]. Therefore, we used estimates from our survey. [4] Negative value for lower bound of 95% CI set to 0. [5] Non-infinite values were removed for calculation of lower bound of 95% CI – an effect size of ≥0 suggests additional funding did not result in more people quitting. [6] These estimates were derived separately and therefore may not exactly match the aggregated age-band estimates. Abbreviations: CI, confidence interval; DLY, discounted life years; ICER, incremental cost-effectiveness ratio; SSS, Stop Smoking Service.

additional-funding/local-stop-smoking-services-and-support-funding-allocations-and-methodology accessed 9 July 2025.

13. Office for National Statistics. Adult smoking habits in the UK: 2024 Newport, United Kingdom: ONS; 2025 [updated 4 November 2025; cited 2026 9 March]. Available from: https://www.ons.gov.uk/peoplepopulationandcommunity/healthandsocialcare/healthandlifeexpectancies/bulletins/adultsmokinghabitsingreatbritain/2024 accessed 9 March 2026.
